# Clinico-Epidemiological Profile of Type II Diabetes Mellitus in India – A Multicentric Institutional-Based Survey

**DOI:** 10.1101/2024.11.06.24316048

**Authors:** Pramila Kalra, Gurinder Mohan, Iadarilang Tiewsoh, K R Raveendra, M Ravi Kiran, Sreejith N Kumar, Sujoy Ghosh, Mala Dharmalingam, Prasanta Kumar Bhattacharya, R Akanth, S Chitra, Kaushik Pandit, Monaliza Lyngdoh, Prabhu Subramani, Pradip Mukhopadhyay, Richa Ghay Thaman, Manish Chandey, B S Ramakrishna, Sivaranjani Holigi, Smitha Jain, R Sundararaman, Srinivas J Vedantha, Veena Sreejith, M D Jamil, Shafiq Rahman, R Mercy Elizabeth, Theertha Sekhar, P R Sreelakshmi, Monika Vempadapu, Aditi Sharma, Reetu Singh, Karan Odedra

## Abstract

**Background:** The epidemic of diabetes mellitus is one of the leading causes of mortality globally. Therefore, the goal of the registry is to create a database on individuals with diabetes mellitus that may be utilized to provide data on the clinico-epidemiological profile of Diabetes Mellitus in the real world.

**Methods:** Data for this registry is captured at seven sites across India recognized by the Biotechnology Industry Research Assistance Council (BIRAC). This observational multi-centric study registered around 25077 Diabetic patients over three years (December 2023).

**Results:** Out of 25077 patients, 12793 (51%) were male and 12284 (49%) females. There were 11443 (46%) rural patients and 13575 (54%) urban patients. Most patients registered were over 50 years old (74.05±2.42). Diabetes was seen as a burden for 46% of individuals and their families. Less than 40% of patients exercised. Over half of the patients had a family history of diabetes. This explains the exponential rise of diabetes mellitus over generations and the significance of preventing it.

**Conclusion:** This registry revealed the impact of the clinico-epidemiological aspects of Diabetes Mellitus on a larger number of samples. Future healthcare planners, researchers, and government officials will benefit from this diabetes registry in developing primary and secondary preventive initiatives that might minimize the rising healthcare burden of diabetes.

## Introduction

According to the IDF (International Diabetes Federation) atlas, in 2021, 537 million persons globally have diabetes, and by 2045, that number could rise to 124.9 million in India alone (1). The burden of T2DM has increased from 1990 to 2019 generally (2). Early onset type 2 diabetes is a growing global health problem in adolescents and young adults, especially in countries with a low-middle and middle socio-demographic index (3). IDF Diabetes Atlas reports, produced annually, present new global epidemiological and diabetes-related impact data and highlight the urgent need for government and policymakers to take action (1).

Patient registries are crucial for medical research and public health surveillance and are a significant resource for understanding, managing, and surveillance of disease. Providing health service providers and planners with data on risk causes and consequences would also aid in monitoring diabetes control programs. The expense and time required for prospective data collection will be minimized using data gathered through registries as an excellent platform for randomized clinical trials. Due to the limited information accessible on Diabetes Mellitus, the Diabetes Registry is exceptionally relevant to the Indian scenario, which provides scope for designing sampling frames for epidemiologic and clinical studies.

In this context, the Department of Biotechnology, Government of India, commissioned a population-based registry project. The current study provides the clinic epidemiological characteristics of the study participants to delineate the critical sociodemographic correlates in the management of Type II Diabetes Mellitus in India.

## Methodology

This hospital-based, multi-centric registry was an observational study collecting data from seven sites across India that were part of the Diabetology Clinical Trial Networks (CTN) and recognized by the Biotechnology Industry Research Assistance Council (BIRAC) (Figure 1). Data were gathered from participants visiting the CTN outpatient departments (OPD) and those with a documented history of Type 2 Diabetes Mellitus. Participants included in the registry were those who provided informed consent and who aged one year and above. Exclusions include non-Indian citizens and those with gestational, drug-induced, or secondary diabetes mellitus. The registry captures a range of information including demographic characteristics, anthropometry, risk factors, complications, coexisting chronic conditions, lifestyle, and concomitant medications using Data Collection Form (DCF). Data are collected from eligible patients through face-to-face interviews using a printed data collection form, following written informed consent/assent from the patients or their parents. An online, restricted-access database was developed to efficiently manage the electronic medical records of diabetes patients (Figure 2).

**Figure 1:**
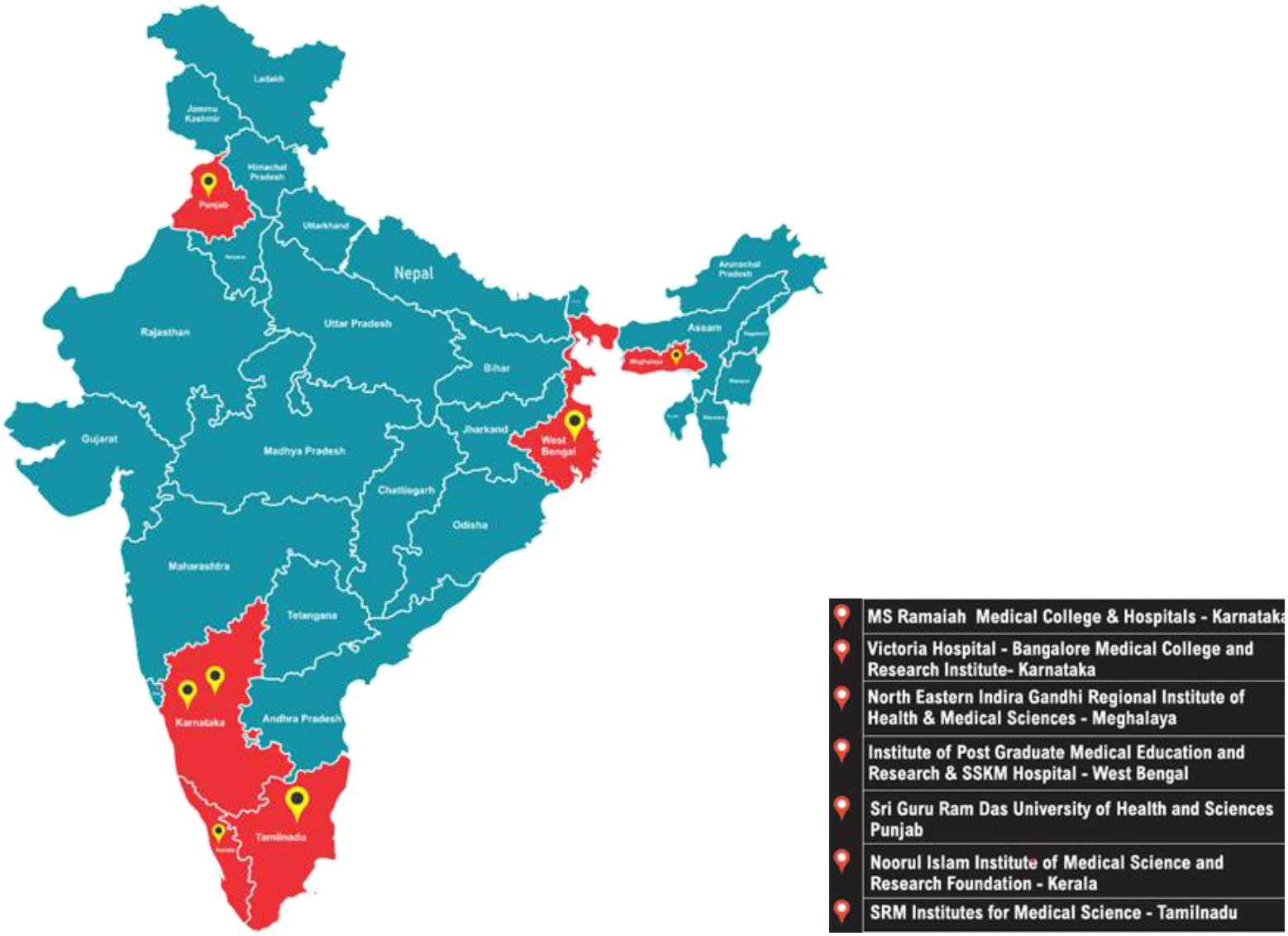
Geographical distribution of Type II Diabetes Mellitus patients based on gender.

**Figure 2:**
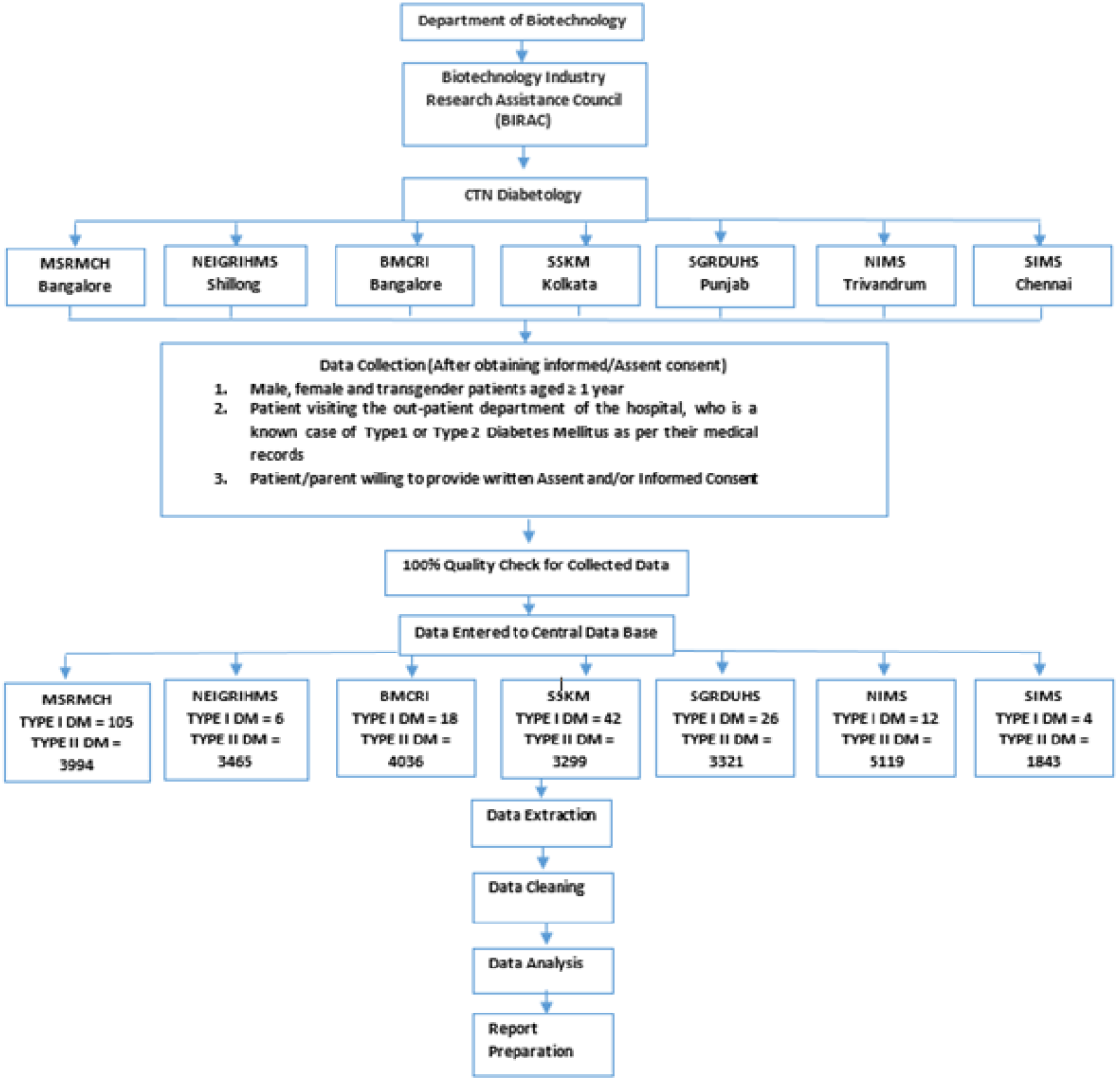
Project Description.

The study was divided into three phases: the preparatory phase, the second phase, and the third phase.

### First Phase

#### Preparatory Phase

Development of protocol and Standard Operating Procedures (SOPs), Validation of the data collection form, Pilot run, Software development for data entry, Ethics Committee approval, Protocol registration under Clinical Trials Registry-India (CTRI) before enrolling the first participant was done in the preparatory phase

### Second Phase

#### Data collection & generate first registry report

Initiation of data collection, Data entry, and verification, and Quality checks

### Third Phase

#### Continuation of data collection & report preparation

Data Collection, Registry report preparation, Manuscript preparation, Publications

## Results

A total of 25077 Type 2 Diabetes Mellitus participants have been enrolled in the registry from all Clinical Trial Network - Diabetology sites, of which 51% of the participants were male and 49% were female. Among the Type 2 Diabetes Mellitus recruited more than 60% of participants were above 50 years of age, also more participants (54.3%) were residing in urban settings.

As per the Kuppuswamy classification of education, occupation and family income of the diabetes mellitus participants enrolled in this study, among male participants 3.8% hold professional degree, 22.9% participants hold graduate degree, and 11.4% participants hold Inter/diploma. More than 50% of the participants have received education equal to or lesser than High school. Almost 9% of participants were illiterate. Among female participants enrolled, 2.8% hold professional degrees, 19.9% participants hold graduate degrees, and 9% participants hold an Inter/diploma. More than 50% of the participants have received education equal to or lesser than High school. Almost 16.5% of participants were illiterate. Among the total male participants recruited 46%, 6% and 11% had professional, semi-professional, and clerical/shop/farming occupations respectively. Almost 10% were under-skilled and semi-skilled workers. Unskilled workers ranged up to only 2% of participants. A total of 55% of female participants were professionals, 5% were semi-professionals, and 8% were clerical/shop/farmers. Almost 8% were skilled and semi-skilled workers. Unskilled workers constitute about 1% of female participants. A total of 23% of participants were unemployed. Family income of 23.3% and 24.9% of male participants ranged from 10002 to 29972 and ≤ 10001 rupees respectively. Among the Type 2 Diabetes Mellitus, 33.9% were not willing to share their income, 28.2% were below 10,001 incomes, and 23.7% were in the 10002-29972 rupees income group. In the females, 34.0% were not willing to share their income, 31.6% had less than 10.001 income, and 21.9% were in the 10,002–29,972 group. Among the males, 33.7% were not willing to share their income, 25.3% were in the 10,002–29,972 group, and 24.9% were below 10,001 incomes. Among Type 2 Diabetes Mellitus patients, 42.4% spend less than 1000 rupees on diabetes treatment, whereas 34.5% spend 1000-5000 rupees, 1.6% spend 5000-10,000 rupees, 0.3% spend more than or equal to 10000 rupees on diabetes treatment. A free supply of medicine is received by 18.6% of diabetes patients enrolled in the study. About 46.0% of patients feel that Diabetes Mellitus is a burden to their family.

Among the patients with Type 2 Diabetes Mellitus, 54.4% were having a family history of Diabetes Mellitus, 29.6% were having a family history of hypertension, and 9.2% were having a family history of CVD. Among the Type 2 Diabetes Mellitus, 99.7% were not doing strengthening, 0.2% were doing strengthening for 15-30 minutes, 0.06% for 30-60 minutes, 98.8% were not doing aerobics, 0.5% were doing aerobics for 15-30 minutes, and 0.5% were doing for 30-60 minutes and 0.2% were doing for more than 1 hour. Among the study patients, 94.9% were not doing flexibility activities, 2.7% were doing it for 15-30 minutes, 1.6% were doing it for 30-60 minutes and 0.8% were doing it for more than 1 hour. Among the study patients, 37.9% were not doing Endurance, 35.9% were doing it for 15-30 minutes, 20.9% for 30-60 minutes, and 5.3% for more than 1 hour.

The mean weight of the study patients was 67.15±13.19; the mean height was 160.02±13.45; the mean Body Mass Index (BMI) was 26.01±4.90; the mean waist circumference was 91.95±12.10. The mean weight and BMI were highest in patients with Glycated hemoglobin (Hb1Ac) more than 7%. Association of age with BMI in Type 2 Diabetes Mellitus shows that the majority of the patients in the category of overweight and obese were in the age groups of 41-50, 51-60, and more than 60 years. The association of age with obesity was significant in Type 2 Diabetes Mellitus (P Value = 0.001 (sig)). The association of gender with BMI in Type 2 Diabetes Mellitus shows that there is a significant association (P Value = 0.001 (sig)). Among the obese category, 64.4% were females, and 35.6% were males. Among the underweight category, 37.1% were females, and 62.9% were males.

Among the patients recruited, the mean random blood sugar (RBS) was 223.69±90.46 mg/dL, the mean fasting blood sugar (FBS) was 164.87±67.40 mg/dL, and the mean postprandial plasma glucose (PPS) was 230.41±98.18. The mean serum creatinine was 1.01±3.03, high-density lipoprotein (HDL) was 45.52±14.90, the mean low-density lipoprotein (LDL) was 104.71±39.38 and the mean very low-density lipoprotein (VLDL) was 35.27±23.70. The mean cholesterol level was 179.37±48.05, and the mean triglycerides level was 169.31±110.35. The patients with Hb1Ac levels higher than 7% had the highest RBS, FBS, PPS, HDL, LDL, Cholesterol and Triglyceride. Based on the association of age groups with levels of Hb1Ac, most patients were in the category of 6.5-6.9% and more than 7% were in age groups of 41-50 years, 51-60 years, and more than 60 years. The association of age with the Level of Hb1Ac was significant in Type 2 Diabetes Mellitus (P Value = 0.001 (sig)). There was also a significant association of gender with the Hb1Ac levels (P Value = 0.001 (sig)). Among HbA1c less than 5.9% category, 542% were males. Among the 6.5-6.9% category, 53.4% were males, and 46.6% were females. Among the more than 7% category, 51.7% were males.

Among the patients with Type 2 Diabetes Mellitus, 7.3% had neuropathy, 6.6% had retinopathy, 2.9% had chronic kidney disease and infections, and 2.0% had nephropathy. Association between gender and complications in Type 2 Diabetes Mellitus shows that the number of patients with complications was higher in males than in females, with a non-significant chi-square association between gender and complications (P Value =0.314 (Non-Sig)). Association between the age groups and the complications showed significance with a P value of 0.001. The percentage of the patients with complications was significantly higher at more than 60 years of age, followed by 51-60 years and 41-50 years. The rate of the patients with complications was lowest in the younger age groups. There was a significant association between the level of Hb1Ac and complications (P Value = 0.001 (sig)). Higher percentages of the patients had complications in those with more than 7% Hb1Ac. The percentage of the patients with complications was less in those with less than 5.9% Hb1Ac and in those with 6-6.4% Hb1Ac.

Among Type 2 Diabetes Mellitus patients, the number of patients who checked for microvascular complications were around 59.7%. 73.7% of the Type 2 Diabetes Mellitus patients have done ECG check in the last 6 months. More than half of the patients (74.8%) follow the diet accurately. More patients (92.1%) follow medication as the General Practitioner advised. Self-measured plasma Glucose (SMPG) is done by only 45.8% of patients, and a small number of patients (3.4%) do CGMS.

## Discussion

According to the IDF (International Diabetes Federation) atlas, in 2021, 537 million persons globally have diabetes, and by 2045, that number could rise to 124.9 million in India alone. In addition, 541 million people are estimated to have impaired glucose tolerance in 2021. It is also estimated that over 6.7 million people aged 20–79 will die from diabetes-related causes in 2021 (1).

In India, the first diabetes registry was set up in Goa as a public-private partnership aimed at population-based disease management. A state-wide campaign over four years has screened and revealed about 44,000 patients with diabetes in the state (4). In Puducherry, a project was started to create a population-based diabetes registry where 2177 patients with diabetes were registered from six Primary Health Centres (5). Similar innovative initiatives under the Changing Diabetes Barometer of Novo Nordisk Education Foundation have targeted states like Bihar and Gujarat. A sum of 12,140 subjects was screened at five primary health centers in Gujarat and found to be 13.1% diabetic and 11.3% prediabetes population (6). Similarly, our study initiated as a Diabetes Registry with 25077 Type 2 Diabetes Mellitus participants enrolled from seven sites across India, provides scope for designing sampling frames for epidemiologic and clinical studies. Our registry, by collecting and sharing quality data, can help in variation analysis, identifying current practices, comparing with existing guidelines for better care, giving proficient feedback, and providing learning opportunities for the health systems.

According to “Clinical and Demographic Profile of Diabetic Patients from Central India - Results from Diabetes Registry,” among 12,434 patients enrolled, 54.95% were below 50 years, and 45.05% were above 50 years. 50.21% were females, and 49.79% were males (7), which is similar to the demographic breakdown of the participants in our study comprising 12793 (51.1%) males and 12284 (48.9%) females. In the ICMR– India Diabetes study, data from 15 states showed that the overall prevalence of diabetes in all 15 states of India was 7·3% (95% CI 7·0-7·5). The prevalence of diabetes varied from 4·3% in Bihar (95% CI 3·7-5·0) to 10·0% (8·7-11·2) in Punjab and was higher in urban areas (11·2%, 10·6-11·8) than in rural areas (5·2%, 4·9-5·4; p<0·0001) (8). In this study, a similar pattern was noticed where 57.9% of the enrolled participants resided in the urban area, and 42.2 % resided in the rural area.

Direct health expenditures due to diabetes are already close to one trillion USD and will exceed this figure by 2030 (1). Type 2 diabetes, which accounts for the majority of diabetic cases, can lead to multiorgan complications, broadly divided into microvascular and macrovascular complications. These complications are a significant cause for increased premature morbidity and mortality among individuals with diabetes, leading to reduced life expectancy and financial and other costs of diabetes leading to a profound economic burden on the Indian healthcare system (9). The age-standardized DALY rate for diabetes increased in India by 39·6% (32·1-46·7) from 1990 to 2016, which was the highest increase among major non-communicable diseases (10). In this study, the occupational status based on the Kuppuswamy scale shows that almost 49% of family heads are unemployed. Among total Type 2 Diabetes Mellitus patients, 34.5% spend 1000-5000 rupees, and 42.4% spend less than 1000 Rs per month on medication. The statistics on the impact of diabetes mellitus show that 1.9% of patients are bedridden, 3.3% have lost their jobs, and 1.4% of patients had lower limb amputation due to diabetes mellitus. About 46% of patients feel that Diabetes Mellitus is a burden to their family.

Evidence suggests that family history by itself is most useful for predicting disease when there are multiple family members affected, the relationship among relatives is close, and the disease is premature, that is, it occurs at younger ages than would be expected (11). The risk of diabetes was 76.3% higher among participants with a family history of diabetes. There is a 41.1% chance of diabetes among those participants whose fathers had diabetes, and a 39.3% chance of diabetes among those participants whose mothers had diabetes (12). In our study 54.4% had a family history of Diabetes Mellitus, 29.6% had a family history of hypertension, and 9.2% had a family history of CVD.

In the ICMRINDIAB study between Oct 18, 2008, and Dec 17, 2020. The overall weighted prevalence of diabetes was 11·4% (95% CI 10·2–12·5; 10151 of 107119 individuals), prediabetes 15·3% (13·9–16·6; 15496 of 107 119 individuals), hypertension 35·5% (33·8–37·3; 35 172 of 111439 individuals), generalized obesity 28·6% (26·9–30·3; 29861 of 110368 individuals), abdominal obesity 39·5% (37·7–41·4; 40121 of 108 665 individuals), and dyslipidaemia 81·2% (77·9–84·5; 14 895 of 18 492 of 25647) (13). A study on “Prevalence of Vascular Complications Among Type 2 Diabetic Patients in a Rural Health Center in South India” included 390 type 2 diabetes patients. The overall prevalence of macrovascular and microvascular complications in our study population was 29.7% and 52.1%, respectively. Among the macrovascular complications, both coronary artery disease (CAD) and peripheral vascular disease (PVD) had a prevalence rate of 15.1%. Among the microvascular complications, peripheral neuropathy (44.9%) had the highest prevalence followed by nephropathy (12.1%) and diabetic foot (7.2%) (14). As per the discovery study program in 38 countries, the median duration of type 2 diabetes was 4.1 years (interquartile range [IQR]: 1.9–7.9 years), and the median glycated hemoglobin (HbA1c) level was 8.0% (IQR: 7.2–9.1%). The crude prevalences of microvascular and macrovascular complications were 18.8% and 12.7%, respectively. Common microvascular complications were peripheral neuropathy (7.7%), chronic kidney disease (5.0%), and albuminuria (4.3%). Common macrovascular complications were coronary artery disease (8.2%), heart failure (3.3%) and stroke (2.2%) (15). In our study, in our study the mean HbA1c value was 9.26 with SD = 3, the most frequent complications observed among Type 2 Diabetes Mellitus are Neuropathy, Diabetic Retinopathy, and Nephropathy, with 7.3%, 6.6%, and 4.9% respectively.

Epidemiological studies have revealed a significant prevalence of undiagnosed diabetes mellitus patients. The absence of screening options, along with the non-specific symptoms of the disease, primarily accounts for a considerable number of patients exhibiting microvascular or macrovascular problems at the initial diagnosis of diabetes. The healthcare delivery system should prioritize diabetes prevention and associated consequences in primary care to alleviate the strain on the tertiary hospital system. It is imperative to amplify diabetes prevention initiatives, fortify care, and improve surveillance within the community to attain the third objective of the Global Sustainable Development Goals. The objective is to decrease premature mortality from non-communicable diseases by one-third by 2030, achievable only by a concentration on diabetes. Our research underscores the necessity for a registry that establishes a database on persons with diabetes mellitus and offers data on the real-world monitoring and management of the condition. This study also assesses the prevalence of comorbidities and the implications of diabetes mellitus. This register can furnish health service providers and planners with valuable insights regarding risk factors and problems. It would assist State health services in ongoing monitoring, identifying individuals at risk of diabetic complications, and bridging the gap between scientific information and clinical outcomes. Registries and other health information technology, including electronic health records, serve as foundational components of health systems. These interventions possess significant potential to enhance decision-making at both individual and community levels. Consequently, the data might be employed to strategize preventive services and enhance resource management.

## Conclusion

Every healthcare system aiming to treat diabetes patients through an internally integrated management program inside their primary care delivery systems must establish a register as a fundamental component. To attain enhanced results for diabetes, a systematic population-based strategy is necessary for controlling all elements of the chronic illness care paradigm. Diabetes registries serve as a crucial epidemiological instrument for monitoring the prevalence and incidence of diabetes, offering a sampling framework for epidemiological and clinical research, supplying health service providers and planners with data on risk factors and complications, and aiding in the comprehensive oversight of diabetes control initiatives. This registry can enhance needs assessment for specialist services, provide a mechanism for complications screening, and help bridge the gap between evidence-based care recommendations and clinical outcomes at the patient level. This register facilitates investigative and analytical tools for gathering accurate and valuable information. It will assist in collecting information, assessing the burden of disease, planning healthcare, allocating resources, developing realistic management guidelines, enhancing clinical practice, and minimizing errors. The registry provides both direct and indirect advantages to society in various domains. This will facilitate the prevention and control of disease patterns.

## Data Availability

All data produced in the present study are available upon reasonable request to the authors

## Limitations of the Study

A few limitations of the study include the dataset does not contain information about hospital admissions, and the limited geographical region of India covered.

## Acknowledgment(s)

The authors gratefully acknowledge the support provided by the Biotechnology Industry Research Assistance Council (BIRAC) and the National Biopharma Mission (NBM). We also appreciate the technical assistance provided by all the staff and management of CTN Diabetology Centre. The authors would like to thank Dr. Nandakumar B S, Professor of Community Medicine at M S Ramaiah Medical College and Hospitals for his valuable insights and feedback on early drafts of this manuscript.

## Tables

**Table 1:** Sociodemographic characteristics of study population.

| Age Group (In Years) | Urban n (%) | Rural n (%) | Total |
| --- | --- | --- | --- |
| 1-10 | 1 (0.01%) | 0 (0%) | 1 (0.01%) |
| 11-20 | 13 (0.1%) | 21 (0.2%) | 34 (0.1%) |
| 21-30 | 185 (1.6%) | 208 (1.5%) | 393 (1.6%) |
| 31-40 | 1147 (10%) | 1081 (7.9%) | 2228 (8.9%) |
| 41-50 | 2942 (25.7%) | 2957 (21.7%) | 5899 (23.5%) |
| 51-60 | 3779 (32.9%) | 4290 (31.5%) | 8069 (32.2%) |
| 61-70 | 2479 (21.7%) | 3476 (25.5%) | 5955 (23.6%) |
| 71-80 | 831 (7.3%) | 1395 (10.3%) | 2226 (8.9%) |
| 81-90 | 80 (0.7%) | 182 (1.3%) | 262 (1.0%) |
| More than 90 | 4 (0.01%) | 6 (0.01%) | 10 (0.01%) |
| **Mean (±Std)** | 54.21±11.36 | 56.24±11.80 | 55.31±11.64 |
| **Gender** |  |  |  |
| **Female** | 5530 (48.2%) | 6754 (49.6%) | 12284 (48.9%) |
| **Male** | 5931 (51.8%) | 6862 (50.4%) | 12793 (51.1%) |
| **Total** | 11461 (100.0%) | 13616 (100%) | 25077 (100%) |

**Table 2:**
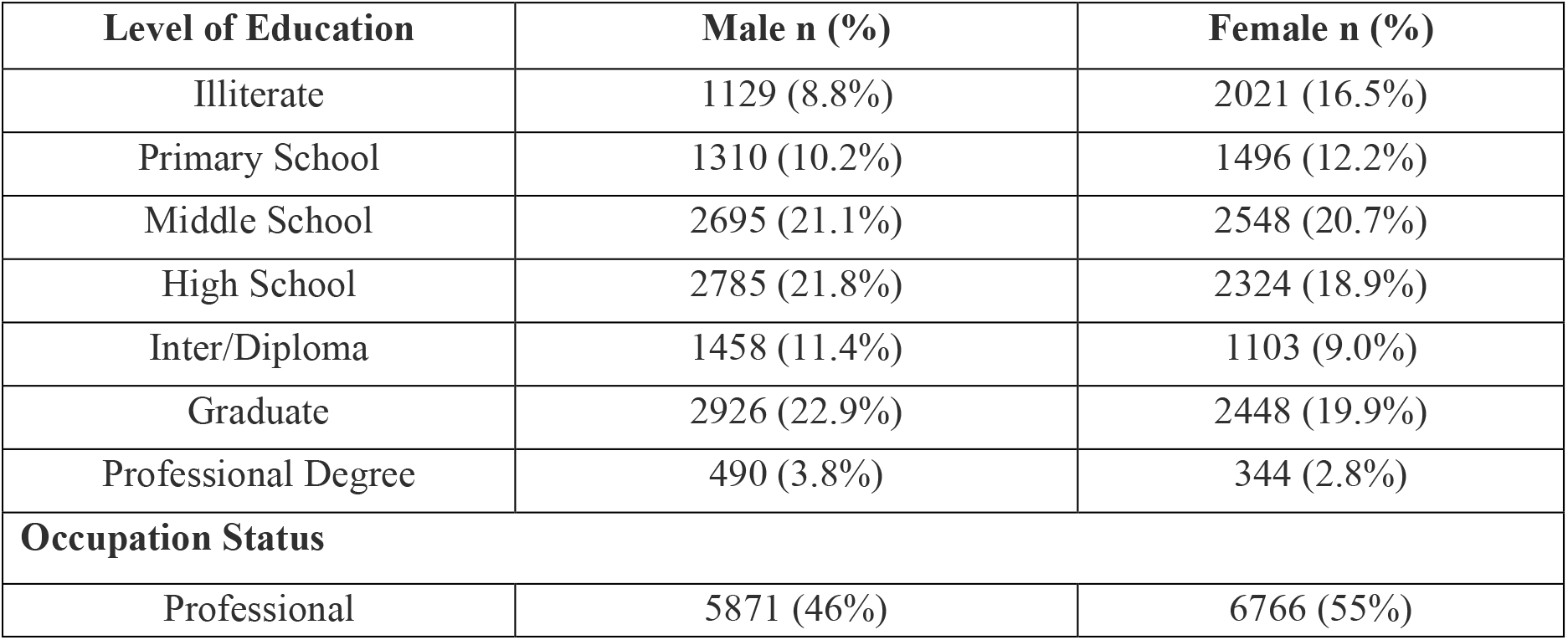
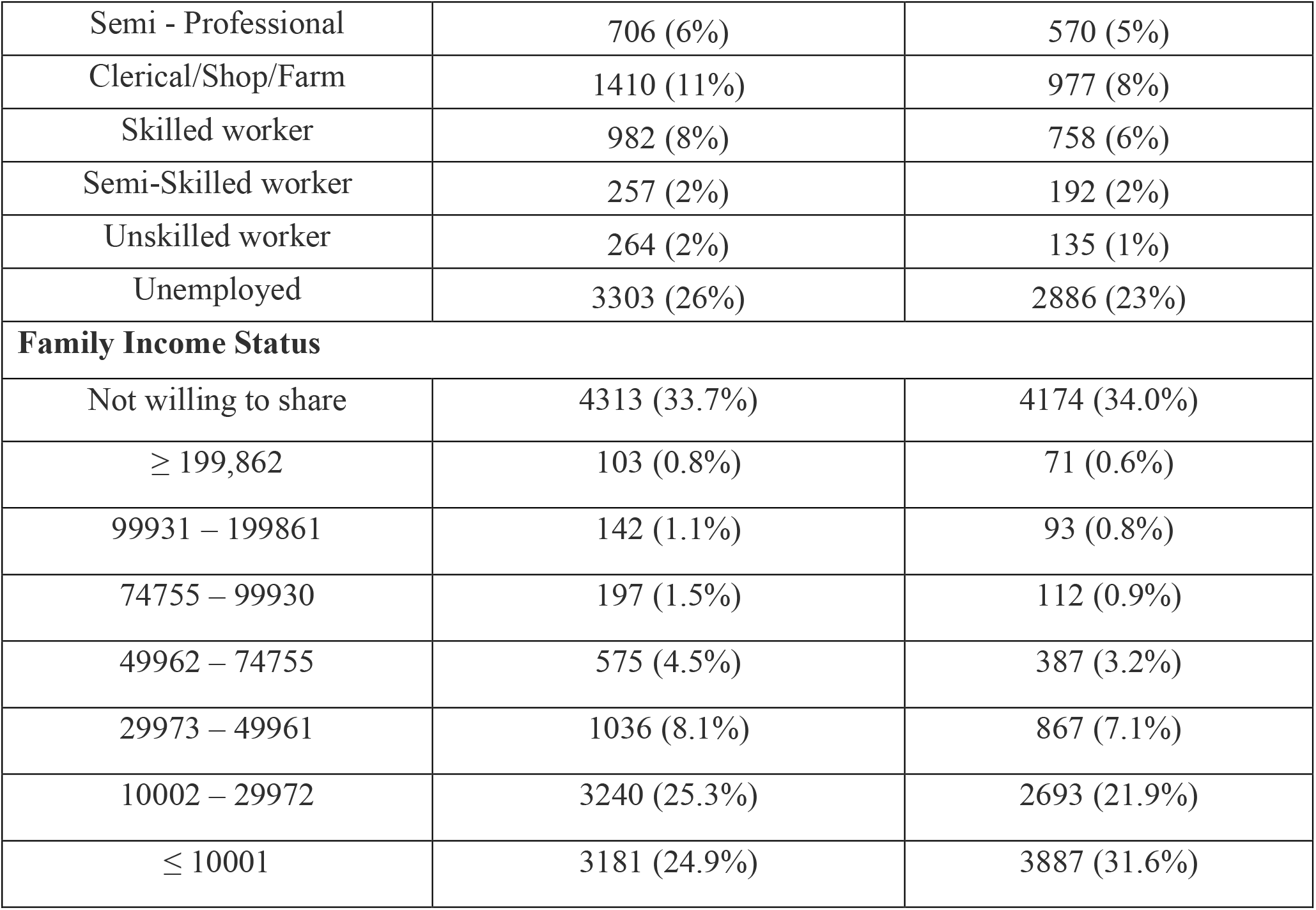
Socioeconomic Characteristics of the study population.

| Level of Education | Male n (%) | Female n (%) |
| --- | --- | --- |
| Illiterate | 1129 (8.8%) | 2021 (16.5%) |
| Primary School | 1310 (10.2%) | 1496 (12.2%) |
| Middle School | 2695 (21.1%) | 2548 (20.7%) |
| High School | 2785 (21.8%) | 2324 (18.9%) |
| Inter/Diploma | 1458 (11.4%) | 1103 (9.0%) |
| Graduate | 2926 (22.9%) | 2448 (19.9%) |
| Professional Degree | 490 (3.8%) | 344 (2.8%) |
| **Occupation Status** |  |  |
| Professional | 5871 (46%) | 6766 (55%) |
| Semi - Professional | 706 (6%) | 570 (5%) |
| Clerical/Shop/Farm | 1410 (11%) | 977 (8%) |
| Skilled worker | 982 (8%) | 758 (6%) |
| Semi-Skilled worker | 257 (2%) | 192 (2%) |
| Unskilled worker | 264 (2%) | 135 (1%) |
| Unemployed | 3303 (26%) | 2886 (23%) |
| **Family Income Status** |  |  |
| Not willing to share | 4313 (33.7%) | 4174 (34.0%) |
| ≥ 199,862 | 103 (0.8%) | 71 (0.6%) |
| 99931 – 199861 | 142 (1.1%) | 93 (0.8%) |
| 74755 – 99930 | 197 (1.5%) | 112 (0.9%) |
| 49962 – 74755 | 575 (4.5%) | 387 (3.2%) |
| 29973 – 49961 | 1036 (8.1%) | 867 (7.1%) |
| 10002 – 29972 | 3240 (25.3%) | 2693 (21.9%) |
| ≤ 10001 | 3181 (24.9%) | 3887 (31.6%) |

**Table 3:** Distribution of physical activity of Type 2 Diabetes Mellitus patients.

| **Physical activity**<br>**(N = 25077)** | **Nil** | **15-30 Minutes** | **30-60 Minutes** | **More than 1**<br>**Hour** |
| --- | --- | --- | --- | --- |
| Strengthening | 24981<br>(99.7%) | 43<br>(0.2%) | 16<br>(0.06%) | 37<br>(0.1%) |
| Aerobics | 24768<br>(98.8%) | 127<br>(0.5%) | 116<br>(0.5%) | 66<br>(0.2%) |
| Flexibility | 23792<br>(94.9%) | 667<br>(2.7%) | 405<br>(1.6%) | 211<br>(0.8%) |
| Endurance | 9497<br>(37.9%) | 9024<br>(35.9%) | 5212<br>(20.9%) | 1340<br>(5.3%) |

